# SARS-CoV-2 seroprevalence in asymptomatic or mild symptomatic people and symptomatic patients with negative PCR results: The hidden perspective in epidemiological reports

**DOI:** 10.1101/2021.07.11.21260336

**Authors:** Nazila Hajiahmadi, Faezeh Mojtahedzadeh, Atefeh Yari, Mahdi Tat, Hoorieh Soleimanjahi, Saeid Amel Jamehdar, Mitra Jafari, Samira Asli, Rohollah Dorostkar, Maryam Nazemipour, Mohammad Ali Mansournia, Taravat Bamdad

## Abstract

**Background:** SARS-CoV-2 has led to the current pandemic of respiratory disease. The reports of confirmed COVID-19 cases based on molecular tests do not completely cover the total number of infected people. These reports do not include the asymptomatic or mildly symptomatic patients and also the patients with false-negative RT-PCR results, while the infection is contagious in all of these conditions.

**Objective:** In this study, we tried to improve our conception of the hidden perspective of SARS-CoV-2 in epidemiological reports.

**Methods:** From May 30 to June 17, 2020, blood samples were collected from two groups of people: asymptomatic or mild symptomatic volunteer participants and severe symptomatic hospitalized patients with negative PCR results. Detection of SARS-CoV-2 antibody was done with ELISA kit targeting N or S proteins.

**Results:** Totally 716 samples from volunteer participants and 81 samples from symptomatic hospitalized patients with negative PCR were evaluated. The test performance-adjusted seroprevalence (95% CI) of SARS-CoV-2 anti-N IgG was 17.3% (8.8%, 25.8%) for volunteers and 25.5% (12.8%, 39.7%) for anti-N and S IgM in hospitalized group. There was an association between high-risk occupations, high-risk behaviors, or symptomatic diseases with positive SARA-Cov-2 N antibody results. Among anti-N positive infected individuals, 49.2% (21.4%, 78.8%) were anti-S positive.

**Conclusion:** The results showed that SARS-COV-2 infection occurs in asymptomatic or mildly symptomatic individuals, but in more than half of them, the produced antibody is not protective. Findings of hospitalized patients also showed that the combination of IgM assay with real-time PCR improves the detection of the disease by more than 25% in negative molecular cases.

## Introduction

The latest member of Coronaviridae, SARS-CoV-2, is the causative agent of the current respiratory disease worldwide (1). The Coronavirus disease 2019 (COVID-19) has caused a spectrum of illnesses which is divided into three levels: asymptomatic or mildly symptomatic disease; the severe disease which requires medical care and disease with serious conditions, and organ failure that requires hospitalization in ICU (2, 3). Due to the similarity of COVID-19 common symptoms such as fever, fatigue, and cough (2, 4) with other respiratory diseases like Influenza (5), the routine diagnosis is based on molecular tests, CT scans, and some hematology factors (6). The official reports of confirmed COVID-19 cases based on molecular tests do not include the asymptomatic or mildly symptomatic patients who don’t refer to medical centers and also the patients with false-negative RT-PCR results, while the infection is contagious in all of these conditions (7, 8). Antibody assay against N protein, as a highly antigenic and widely expressed protein during infection, can retrospectively detect the status of the infection in the community, even in asymptomatic cases, and also helps diagnose the disease in case of false-negative molecular tests. However, since N does not induce neutralizing antibodies, the presence of this antibody does not confirm protectivity. Due to the presence of neutralizing epitopes in S protein, anti-S antibodies can to some extent represent the protection of individuals against re-infection. To improve our conception about SARS-CoV-2 seroprevalence, in the current study, N and S antibody status were evaluated in asymptomatic or mildly symptomatic volunteer participants and severe symptomatic hospitalized patients with negative PCR results. The role of job status and social behaviors in antibody prevalence was also evaluated.

## Material and methods

### Samples

From May 30 to June 30, 2020, peripheral venous blood samples were collected from two groups of people from Tehran and Mashhad cities: (1) asymptomatic, mildly symptomatic, or symptomatic volunteer participants who have not referred to medical centers due to COVID-19 symptoms and (2) severe symptomatic hospitalized patients with negative PCR results. Written informed consents were obtained before sampling and each volunteer was asked to answer a questionnaire form including demographic data such as age, gender, occupation, high-risk behaviors, and history of symptoms compatible with COVID-19 from February 2020. Occupation data were divided into low-risk and high-risk jobs (like hospital clinical and nonclinical staffs, taxi drivers, and jobs in crowded places like peddlers). The volunteer participants with a history of traveling, use of public transportation, contact with symptomatic patients, or working at high-risk places were considered as high-risk behavior volunteers. A history of symptoms compatible with COVID-19 from February 2020 was divided into asymptomatic, mild symptomatic (fatigue, runny nose, sneezing, nausea, diarrhea), symptomatic (fever, cough, shortness of breath), and severe symptoms that need hospitalization. Sera were separated immediately and stored at -20°C until testing. Symptomatic volunteers at sampling time were excluded from volunteer group. Ethics Committee of Tarbiat Modares University (IR.MODARES.REC 1399.009) and Baqiyatallah University of Medical Sciences (IR.BMSU.REC.1399.299) approved the study.

### Serological assessment

Detection of SARS-CoV-2 antibody was done with ELISA kit targeting N or S protein (PISHTAZTEB-Iran) according to manufacturer’s protocol. Anti-N IgG SARS-CoV-2 antibody ELISA kit with 94.1% sensitivity and 98.3% specificity and anti-S IgG kit with 85.3% sensitivity and 99.01 % specificity was used for samples of volunteer participants. IgM SARS-CoV-2 antibody ELISA kit coated with N and S proteins with 79.4% sensitivity and 97.3% specificity was used for samples of hospitalized patients with negative PCR results. The optical densities that were greater than the cut off value were considered positive.

### Statistical analysis

The prevalence proportions of anti_N IgG, anti_S IgG, and anti N and S IgM seropositivity with 95% confidence intervals (CIs) were estimated, adjusting for clustering by location, through cluster robust standard errors (9-12). The prevalence estimates were adjusted for the test performance i.e., sensitivity (Se) and specificity (Sp) using a Monte Carlo bias analysis with 100,000 samples based on the following formula: TP = (AP + Sp - 1) / (Se + Sp - 1) where TP denotes true prevalence and AP denotes apparent prevalence (13, 14). We used betta distribution for sensitivity and specificity and set α = the number of true positives and β = the number of false negatives, reported by the manufacturer (PISHTAZTEB-Iran), so that the mean and variance of betta distribution approximately equal the mean and variance of sensitivity or specificity estimates. The apparent prevalence was drawn from a Normal distribution with mean and standard deviation equal to prevalence estimate and its standard error, respectively. We derived the point estimates and 95% simulation intervals (for simplicity, called confidence intervals in this paper) using the median and 2.5^th^ and 97.5^th^ percentiles of Monte Carlo distribution (15). Multiple logistic regression was used to obtain the adjusted odds ratios (ORs) with 95% CIs between variables and anti_N IgG seropositivity (16-18). Paired t-test was used for comparing anti-N and anti-S titers on the logarithmic scale, and the results were presented as geometric mean ratio with 95% CI. All analyses were performed in Stata version 14 (Stata, https://www.stata.com/).

## Results

Totally 797 serum samples were collected. Seven hundred and sixteen samples were from asymptomatic or symptomatic volunteer participants who did not seek medical care because of COVID-19 symptoms, and 81 samples were from symptomatic hospitalized patients that their SARS-CoV-2 molecular results were negative. The median age was 37.4 years (range: 5-92 years) in volunteer participants and 56.07 years (range:20-92 years) in symptomatic hospitalized patients. Sex distribution in volunteer participants and symptomatic hospitalized patients were comprised 50.7% and 50% females respectively. Almost 50% of volunteers had high-risk jobs, that 39% of them were clinical or non-clinical hospital staff. The behavior of 70.8% of volunteer participants was considered as high-risk. The characteristics of samples are summarized in Table I.

**Table I:**
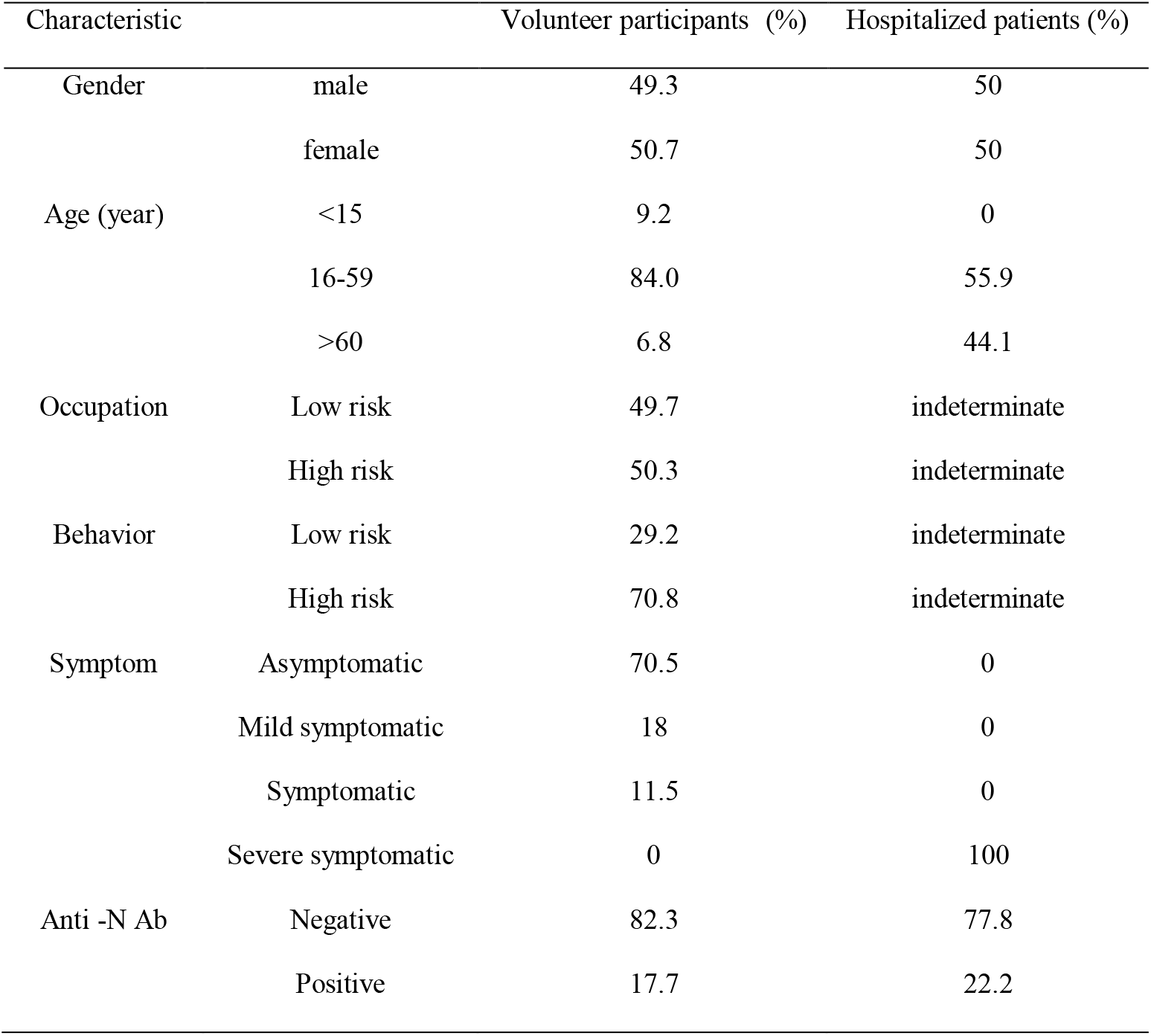
Characteristics of volunteer participants and hospitalized patients with negative PCR results.

| Characteristic |  | Volunteer participants (%) | Hospitalized patients (%) |
| --- | --- | --- | --- |
| Gender | male | 49.3 | 50 |
|  | female | 50.7 | 50 |
| Age (year) | <15 | 9.2 | 0 |
|  | 16-59 | 84.0 | 55.9 |
|  | >60 | 6.8 | 44.1 |
| Occupation | Low risk | 49.7 | indeterminate |
|  | High risk | 50.3 | indeterminate |
| Behavior | Low risk | 29.2 | indeterminate |
|  | High risk | 70.8 | indeterminate |
| Symptom | Asymptomatic | 70.5 | 0 |
|  | Mild symptomatic | 18 | 0 |
|  | Symptomatic | 11.5 | 0 |
|  | Severe symptomatic | 0 | 100 |
| Anti -N Ab | Negative | 82.3 | 77.8 |
|  | Positive | 17.7 | 22.2 |

SARS-CoV-2 positive antibody prevalence proportions (95% CI) in volunteer participants and hospitalized patients with negative SARS-CoV-2 PCR were 17.7% (10.4%, 25.1%) and 22.2% (13.1%, 31.3%), respectively; After adjustment for test performance, they were 17.3% (8.8%, 25.8%) and 25.5% (12.8%, 39.7%). About 70% of volunteers didn’t have any symptoms related to SARS-CoV-2, while 13.7% of them have positive COVID-19 antibody results.

Subjects aged between 15 and 60 years (compared to those less than 15 years old) and men as well as those with high risk occupations (hospital workers and taxi drivers), high risk behaviors, and symptomatic diseases are at a greater risk of positive anti-N SARS-CoV-2 IgG (Table II).

**Table II:** Adjusted associations between characteristics and COVID-19 Ab results in volunteer participants.

| Characteristic |  | COVID-19<br>Anti N IgG<sup>-</sup> (%) | COVID-19<br>Anti N IgG<sup>+</sup> (%) | OR* (95% CI) | p-value |
| --- | --- | --- | --- | --- | --- |
| Gender | female | 83.6 | 16.4 | - |  |
|  | male | 82.2 | 17.8 | 1.39 (1.2, 1.89) | 0.039 |
| Age (year) | <15 | 84.1 | 15.9 | - |  |
|  | 16-59 | 81.7 | 18.3 | 1.30 (1.07, 1.56) | 0.007 |
|  | >60 | 87.2 | 12.8 | 1.19 (0.90, 1.57) | 0.23 |
| Occupation | Low risk | 88 | 12 | - | <0.001 |
|  | High risk | 78 | 22 | 1.66 (1.57, 1.76) |  |
| Behavior | Low risk | 89.5 | 10.5 | - | <0.001 |
|  | High risk | 80.5 | 19.5 | 1.29 (1.24, 1.35) |  |
| Symptom | Asymptomatic | 86.3 | 13.7 | - |  |
|  | Mild symptomatic | 80.6 | 19.4 | 1.40 (0.73, 2.67) | 0.31 |
|  | Symptomatic | 61 | 39 | 4.00 (2.25, 7.10) | <0.001 |

\*ORs are adjusted for other variables in the table

SARS-Cov-2 S antibody was evaluated in SARS-Cov-2 N positive antibody volunteer participants (Fig I). Among N positive subjects, the percentage of S positive individuals was 42.5% (19.3%, 65.7%); the performance-adjusted prevalence was 49.2% (21.4%, 78.8%). In other words, the results showed that in 50.8% (21.2%, 78.6%) of positive SARS-CoV-2 N antibody volunteer participants, S antibody which is more correlated with neutralizing antibodies is negative. Analysis of N and S antibody titers (OD values) showed that geometric mean in anti-N titer was 3.86, while for that of anti-S was 0.72. The geometric mean ratio with 95% CI was 5.36 (4.25, 6.75); P<0.001.

**Fig I.**
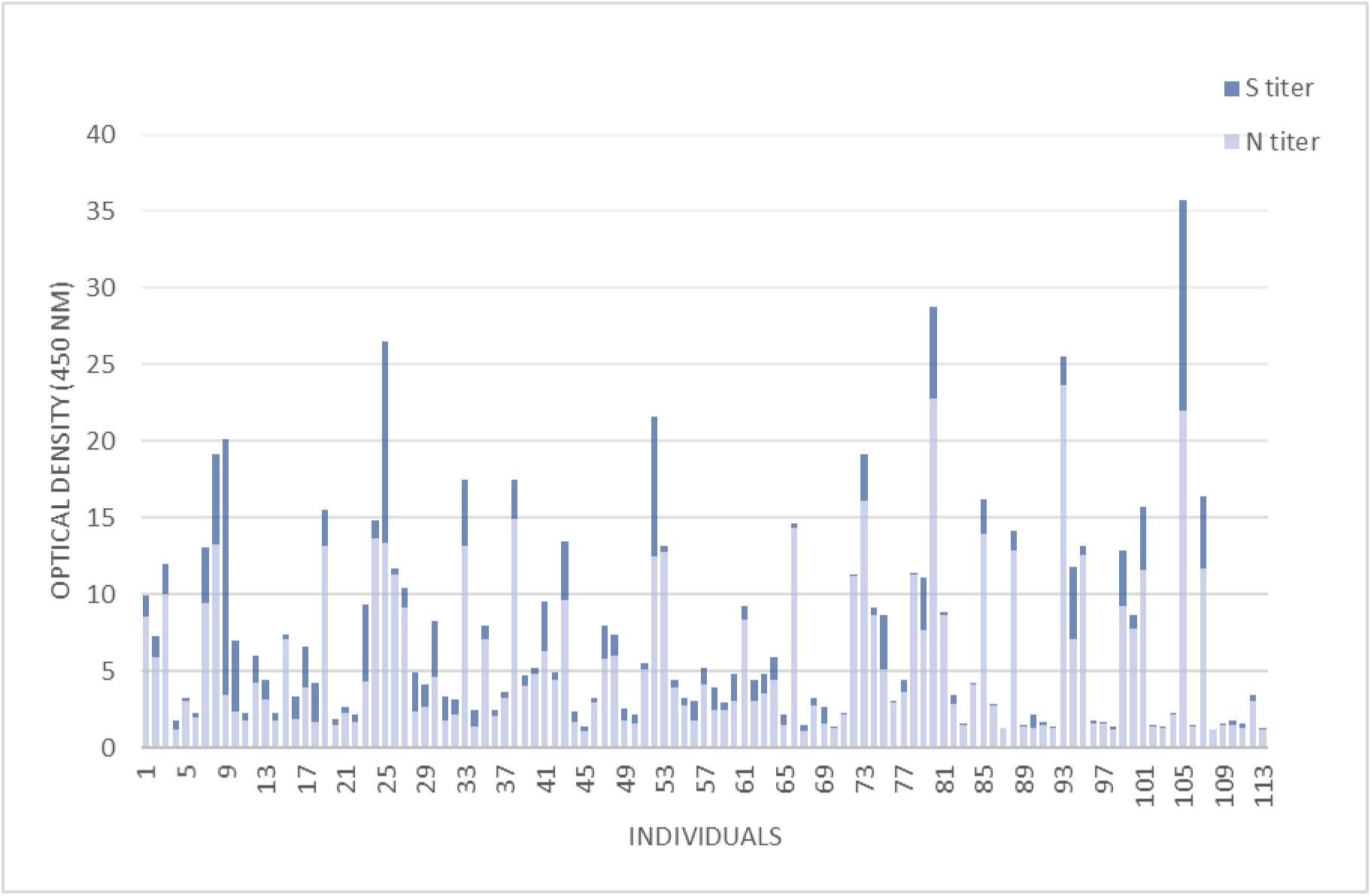
Compairing SARS-COV-2 anti-N and anti-S IgG level (OD values) in each seropositive volunteer.

## Discussion

COVID-19 patients with false-negative PCR results as well as asymptomatic or mildly symptomatic SARS-CoV-2 infected people are not included in official reports while they may contribute to virus spread (7, 8). They also can play a role in breaking the chain of virus transmission as immune individuals. The present study showed SARS-CoV-2 antibody situation in volunteer participants with mild or no COVID-19 symptoms and also in hospitalized patients with negative SARS-CoV-2 PCR results. The obtained data is consistent with the reported seroprevalence in other regions of Iran in a relatively similar period of time (10,11). This indicates that a number of people in the community are infected with the virus without symptoms and confirmed PCR test. However, these individuals may not be protected against the virus because of not having neutralizing antibodies. So even assuming that the anti-S immune response in infected people is long-lasting and protective against re-infection, herd immunity strategy is not easily achievable without mass vaccination (3, 19, 20).

This study showed that there are associations between positive SARS-CoV-2 antibody results and high-risk behavior such as traveling, use of public transportation, contact with symptomatic patients, and working at high-risk places (like hospital staff, taxi drivers, and working in crowded places like peddlers). This finding is consistent with the study that showed there is a relation between the diagnosis of asymptomatic COVID-19 infected people and virus-contaminated environments (21). So continuous monitoring, restriction of the passage of people in crowded areas, and disinfecting environments with risk of contamination along with mass vaccination in a short period of time can be useful in decreasing patients and prevention of disease spread.

Consistent with a cross-sectional study that has investigated SARS-CoV-2 antibodies in asymptomatic hospital staff (22), this survey demonstrated that there is an association between positive SARS-CoV-2 antibody results and high-risk occupation. A high percentage of people with high-risk jobs in this study were clinical or non-clinical hospital staff. This means that better protection protocols must be done in medical centers to reduce the emergence of new cases (23). Vaccination of high-risk groups, evaluating the production of neutralizing antibodies, and its persistence after vaccination should be one of the priorities of the health system.

As expected, the data of this survey indicated the existence of false-negative results in molecular tests. Simultaneous molecular and serological COVID-19 tests lead to the decrease of false-negative molecular test results. Accordingly, it can help to timely isolation of COVID-19 patients and decrease the virus spread.

This study had several limitations such as the limited number of participants in each study group which reduced the chance of randomized sampling. In addition, collecting the information of COVID-19 related symptoms was through self-reporting. Besides, the newly infected people without seroconversion have been missed as well as probable false-positive results due to the cross-reactivity of the serological test with other coronaviruses.

In conclusion, the obtained results revealed the existence of virus spreading risk among the cases without confirmed symptoms or molecular tests, but these infected individuals may not produce protective antibodies. The results of this study reflect the Ab status of the population only at the sampling time and due to the variability of recent epidemic conditions in the community, continuous studies in this field are necessary to clarify the situation and extent of COVID-19 in society.

## Data Availability

All the data mentioned in the article are available.

## Ethical statements

Ethics Committee of Tarbiat Modares University (IR.MODARES.REC 1399.009) and Baqiyatallah University of Medical Sciences (IR.BMSU.REC.1399.299) approved the study. Written informed consents were obtained before sampling.

## Acknowledgements

This study was supported financially by a grant of Research and Technology (National Institute of Genetic Engineering and Biotechnology) and school of medical sciences of Tarbiat Modares University. We would like to thank Mashhad and Baqiyatallah University of Medical Sciences for providing valuable specimens and also the Health Center of Tarbiat Modares University for providing the place for volunteer sampling.

